# A multidisciplinary assessment of ChatGPT’s knowledge of amyloidosis

**DOI:** 10.1101/2023.07.17.23292780

**Authors:** Ryan C. King, Jamil S. Samaan, Yee Hui Yeo, David C. Kunkel, Ali A. Habib, Roxana Ghashghaei

## Abstract

Amyloidosis is a rare, multisystem disease with several subtypes including AA (secondary), AL (amyloid light chain), and ATTR (transthyretin amyloidosis). In addition to variable symptoms and multidisciplinary management, amyloidosis being a rare disease further contributes to patients being at risk for decreased health literacy regarding their condition. Increased access to education materials containing simple, plain language may bridge literacy gaps and improve outcomes for patients with rare diseases such as amyloidosis. The large language model (LLM), Chat Generative Pre-Trained Transformer (ChatGPT), may be a powerful tool for improving the availability of accurate and easy to understand education materials. Amyloidosis-related questions from cardiology, gastroenterology, and neurology were sourced from esteemed medical societies and institutions along with amyloidosis Facebook support groups and inputted into ChatGPT-3.5 and GPT-4. Answers were graded on 4-point scale with both models responding to the majority of questions with either “comprehensive” or “correct but inadequate” answers with only 1 (1.2%) answer by GPT-3.5 graded as “completely inaccurate”. When assessing reproducibility, GPT-3.5 scored reliably on more than 83.3% of responses, while GPT-4 produced above 98.2% consistent answers. Our findings show that ChatGPT can potentially serve as a supplemental tool in disseminating vital health education to patients living with amyloidosis.

## Introduction

Amyloidosis is a chronic multisystem disease that comprises several subtypes including AA (secondary), AL (amyloid light chain), and ATTR (transthyretin amyloidosis), with the latter two being the most common but often underdiagnosed [1]. AL amyloidosis is diagnosed in roughly 2,500 to 5,000 individuals annually in the United States (US), while the exact incidence of ATTR and AA remain unknown due to challenges and delays in diagnosis. This uncertainty is attributed to the diverse range of symptoms affecting this patient population who often have varying presentations affecting multiple organ systems [2, 3]. Diagnosing and caring for patients living with amyloidosis relies on effective multidisciplinary collaboration between specialists in fields including but not limited to cardiology, gastroenterology, and neurology [4].

In addition to the complex nature of symptoms and management, Amyloidosis is also considered a rare disease, a designation further contributing to this patient population being at risk for decreased health literacy regarding their condition. A notable scarcity of patient education materials (PEMs) exists for rare diseases compared to common ones, with nearly a tenfold difference which has previously been shown to adversely affect health outcomes [5]. The Centers for Disease Control and Prevention (CDC) posits that improved health literacy could prevent up to one million hospitalizations annually and save $25 billion in total healthcare costs [6]. Increased access to education materials containing simple, plain language is a promising strategy to help bridge literacy gaps and improve outcomes especially for patients with rare diseases such as amyloidosis.

Artificial intelligence (AI), an emerging technology, may be a powerful tool for improving the availability of accurate and easy to understand information for rare and complex diseases like amyloidosis. Chat Generative Pre-Trained Transformer (ChatGPT), an AI-driven large language model (LLM) released in late 2022, has gained widespread adoption, attracting 1.8 billion users per month. [7]. Unlike traditional search engines, which return web page listings, ChatGPT generates human-like text in a structured, conversational format through an intuitive user interface. This is achieved via reinforcement learning from human feedback (RLHF), wherein the model’s responses are refined through feedback loops to optimize performance [8]. With ongoing improvement and training on an extensive dataset spanning diverse topics including medicine, ChatGPT’s accuracy and reliability in answering questions are expected to increase. In March of this year GPT-4.0, the predecessor to the original GPT-3.5, was released and has demonstrated its superior performance in answering clinical questions [9, 10]. This rapid improvement in performance over a short period of time makes large language models a potential asset for both patients and healthcare providers seeking information on diseases like amyloidosis.

As with any emerging technology, rigorous evaluation is essential to ensure its efficacy and safety. It is imperative to evaluate the capabilities and limitations of these models during their nascent stages to detect knowledge gaps before their broad adoption by patients and providers. Earlier studies have demonstrated ChatGPT’s impressive accuracy and reliability in answering clinical questions related to coronary artery disease, cirrhosis, and bariatric surgery [11, 12, 13]. This study aims to build upon previous literature by employing a multidisciplinary approach to assessing ChatGPT’s 1) accuracy in answering questions related to amyloidosis, particularly concerning cardiology, gastroenterology, and neurology; 2) reproducibility of responses; and 3) performance improvement of GPT-4 compared to GPT-3.5.

## Materials and Methods

A total of 98 amyloidosis-related questions were sourced from esteemed medical societies and institutions and inputted into ChatGPT. Questions from amyloidosis Facebook support groups were incorporated for a more comprehensive patient perspective. Of these, 56 addressed general amyloidosis topics, while 42 were specific to cardiology (12), gastroenterology (15), and neurology (15). Each question was inputted twice into both GPT-3.5 and GPT-4, yielding two distinct responses per question for each model. Neurology-related questions were only inputted into GPT-4. Responses were assessed on a scale: 1) Comprehensive, 2) Correct but inadequate, 3) some correct and some incorrect 4) Completely incorrect. Reproducibility was evaluated by categorizing responses into those containing either no incorrect information (grades 1 and 2) or those with some or completely incorrect information (grades 3 and 4). Two independent reviewers, board-certified in cardiology and gastroenterology with expertise in amyloidosis, assessed general questions and questions in their respective specialties. Discrepancies in general question grading were resolved through discussion to reach a consensus. An additional reviewer, board-certified in neurology and specializing in amyloidosis, graded the neurology-specific responses for GPT-4. Microsoft Excel (version 16.68) was used to conduct the statistical analysis.

## Results

Both ChatGPT models responded to the majority of questions with either “comprehensive” or “correct but inadequate” answers (**Table 1**). GPT-4 demonstrated comprehensive responses more frequently than GPT-3.5 for general questions (94.6% vs 85.7%) and gastroenterology (60.0% vs 53.3%). For specialty-specific responses, Cardiology was graded the highest for both models with both receiving 83.3% comprehensive scores. There was a total of 8 (9.6%) responses for GPT-3.5 containing “some correct and some incorrect” information compared to 5 (5.1%) for GPT-4. One gastroenterology question answered by GPT-3.5 received the only (1.2%) “completely incorrect” grade in response to the evidence for using supplements like probiotics and digestive enzymes to enhance digestion. When assessing reproducibility of specialty-specific responses, GPT-3.5 scored reliably 96.4% on general questions, 83.3% for cardiology, and 93.3% for gastroenterology (**Table 2**). GPT-4 produced reproducible responses for 98.2% of general responses and 100% of responses for all specialties. A distinction between models was observed in responses to the prevalence and presentation of systemic multiorgan amyloidosis. GPT-3.5 gave a broad overview omitting the ATTR subtype, whereas GPT-4 included details on systemic involvement for each subtype.

**Table 1.** Accuracy of ChatGPT for answers to amyloidosis related questions. Accuracy grading was based on a scale of 1 = comprehensive, 2 = correct but inadequate, 3 = some correct and some incorrect, and 4 = completely incorrect. Differences in grading between reviewers were resolved through discussion to arrive at a final score for a given answer.

|  | GPT-3.5 | GPT-4 |
| --- | --- | --- |
| General, Total (N=56) |  |  |
| 1 | 48 (85.7%) | 53 (94.6%) |
| 2 | 4 (7.1%) | 3 (5.4%) |
| 3 | 4(7.1%) | 0 (0.0%) |
| 4 | 0 (0.0%) | 0 (0.0%) |
| Differences in grading | 15 (25.9%) | 17 (29.3%) |
| Cardiology (N=12) |  |  |
| 1 | 10 (83.3%) | 10 (83.3%) |
| 2 | 0 (0.0%) | 2 (16.7%) |
| 3 | 2 (16.7%) | 0 (0.0%) |
| 4 | 0 (0.0%) | 0 (0.0%) |
| GI (N=15) |  |  |
| 1 | 8 (53.3%) | 9 (60%) |
| 2 | 4 (26.7%) | 3 (20%) |
| 3 | 2 (13.3%) | 3 (20%) |
| 4 | 1 (6.7%) | 0 (0.0%) |
| Neurology (N=15) |  |  |
| 1 | - | 10 (66.7%) |
| 2 | - | 3 (20%) |
| 3 | - | 2 (13.3%) |
| 4 | - | 0 |

**Table 2.** Reproducibility of ChatGPT answers for amyloidosis related answers. Reproducibility was graded based on responses containing correct or incorrect information with grouping of scores of 1 and 2 (comprehensive; correct but inadequate) vs 3 and 4 (some correct and some incorrect; completely incorrect) together.

|  | GPT-3.5 | GPT-4 |
| --- | --- | --- |
| General (N=56) | 54 (96.4%) | 55 (98.2%) |
| Cardiology, (N=12) | 10 (83.3%) | 12 (100%) |
| GI (N=15) | 14 (93.3%) | 15 (100%) |
| Neurology (N=15) | - | 15 (100%) |

## Discussion

### Large language models are an emerging technology

There is a growing body of literature examining ChatGPT’s knowledge related to common and prevalent health conditions, but studies evaluating its performance for rare diseases are limited. In this study, we used an interdisciplinary panel of experts in amyloidosis from cardiology, gastroenterology, and neurology to evaluate the accuracy and reliability of GPT-4 and GPT-3.5 in answering amyloidosis related questions. Both models produced comprehensive responses to over 85% of general questions, with GPT-4 outperforming. GPT-3.5 produced responses containing inaccurate information slightly more often than GPT-4 (10.8% vs 5.1%) and provided the only “completely inaccurate” response of the study for one gastroenterology question. For cardiology questions, both models surpassed their performance in gastroenterology and neurology. This higher proficiency may stem from the prevalence of cardiac manifestations in amyloidosis and the models’ possible enhanced exposure to relevant data during training. With over 83.3% reproducibility for GPT-3.5 and over 98.2% for GPT-4, GPT’s high reliability and accuracy in this study further bolsters its prospective utility in aiding patients and providers to improve amyloidosis outcomes through enhanced patient health education. While its performance is impressive, we stress the role of these large language models as adjunct rather than replacement of care provided by a team of licensed healthcare professionals.

Previous studies have also shown ChatGPT’s commendable performance in accuracy and reliability concerning cardiovascular disease prevention queries, with the majority of responses deemed appropriate and dependable [11]. In more intricate scenarios encompassing clinical vignettes describing atrial fibrillation, congenital heart disease, heart failure, and cholesterol levels, ChatGPT’s responses were assessed as predominantly reliable, valuable for patients, and crucially, not hazardous. Impressively, many of these responses were favored over those generated by a standard Google search [14].

Yeo et al. (2023) demonstrated that GPT-3.5 accurately answered over 75% of questions related to basic knowledge and 66% of diagnostic questions concerning cirrhosis and hepatocellular carcinoma [12]. Notably, in a follow up study on cirrhosis the authors demonstrated that GPT-4 significantly improved over GPT-3.5, providing superior performance and rectifying errors made by its predecessor [9]. Another improvement by GPT-4 over GPT-3.5 was seen in a prior study examining GPT’s performance on the United Kingdom neurology licensing exam revealed that GPT-3.5 did not achieve a passing score, while GPT-4 passed comfortably [15]. Additionally, this study highlighted GPT’s generalizability in providing accurate information based on medical guidelines from regions outside the US. These findings indicate that both ChatGPT models can reliably provide accurate information on a wide range of clinical queries and that their capabilities are continually evolving. The discrepancy in accuracy and minimal improvement between models seen in our study compared to prior work may be due to the burgeoning data regarding amyloidosis of the GI tract and its rare nature.

Given the relatively recent release of ChatGPT, data testing the Large Language Model’s (LLM) clinical accuracy concerning rare diseases such as amyloidosis is scarce. Mehnan et al. have demonstrated remarkable diagnostic precision for both common and rare diseases, with GPT-4 notably outperforming GPT-3.5. Interestingly, the authors suggested that GPT’s responses weren’t merely a reiteration of existing online content, attributing its capacity to suggest a comprehensive differential diagnosis to an understanding of its rationale [10]. This assessment of GPT displaying a near human-like understanding may be due to its increased training on human dialogue through RLHF and increased parameters included in the GPT-4 dataset [16].

Although the use of ChatGPT should always complement a healthcare provider’s guidance, this emergent technology could prove beneficial for both patients and providers when applied to rare diseases like amyloidosis in the future but in its current state the model requires further testing of its limitations. The LLM has the potential to simplify and increase accessibility of patient education materials (PEMs), thereby fostering health education-driven empowerment through conversational interactions. With the continuous evolution of ChatGPT’s capabilities and the easy-to-use interface, it is expected that its user base will correspondingly expand. The prospect of expedited diagnoses and establishing care with a multidisciplinary medical team in a timelier manner could be additional impacts of ChatGPT and ultimately improve outcomes for patients living with amyloidosis.

### Strengths and Limitations

This study is among the first in employing a multidisciplinary approach to evaluate ChatGPT’s knowledge of amyloidosis, leveraging expertise from physicians across various specialties. This holistic approach enabled a thorough assessment of ChatGPT’s abilities in addressing clinical queries related to amyloidosis, a rare disease necessitating advancements in health education, diagnostics, and management for improved patient outcomes. Moreover, this is the inaugural study to scrutinize GPT’s performance in the context of amyloidosis.

ChatGPT’s limitations encompass the undisclosed nature of its primary training dataset and the absence of citations in its responses to medical queries. Incorporating references to reputable sources, such as esteemed medical society websites or peer-reviewed studies, would enhance clinical relevance and reliability. Additionally, ChatGPT occasionally exhibits a phenomenon termed ‘hallucinations,’ wherein it generates confident but entirely spurious responses [8].

While this study’s multidisciplinary approach was comprehensive, it relied on a single physician reviewer from each specialty. Future research could bolster validity by engaging multiple reviewers within each specialty to minimize the potential for subjective bias. It would also be beneficial to include physicians specializing in hematology, oncology, and nephrology as reviewers due to the integral involvement of those specialties in caring for patients with amyloidosis.

## Conclusion

ChatGPT delivered accurate and reliable responses to amyloidosis related questions across general and specialty-specific queries. ChatGPT can potentially serve as a supplemental tool in disseminating vital health education to patients and assisting providers in addressing diagnostic challenges of this rare disease. However, the presence of some incorrect responses underscores the necessity of utilizing this technology alongside a team of licensed healthcare professionals.

## Data Availability

All data produced in the present study are available upon reasonable request to the authors.

## Acknowledgements

ChatGPT-4 was used in the editing process of this manuscript.

## Funding Statement

There was no funding obtained for this study.

## Declaration of interest statement

The authors have no disclosures or conflicts of interest to declare.

## Notes

### Competing Interest Statement

The authors have declared no competing interest.

### Funding Statement

This study did not receive any funding.

